## Supplement 2 - Glossary for "Spatial Dynamics of Malaria Transmission"

#### Contents

The following has a list all of the parameters, variables, and terms defined and used in the paper. First, we define **reproductive numbers** and the **abbreviations**. Next, the terms are organized alphabetically in sets:

- Calligraphic Script, $\mathcal{A} - \mathcal{Z}$
- Upper Case Roman, $A - Z$
- Lower Case Roman, $a - z$
- Upper Case Greek, $A - \Omega$
- Lower Case Greek, $\alpha - \omega$

#### Reproductive Numbers

Here we summarize the notation used in the text to define reproductive numbers. For a longer discussion, see Supplement 8.

$R_0$ - **The Basic Reproductive Number** is a scalar. Here, we have defined $R_0$ as the *spectral radius* of one of the parasite's next generation matrices, either $\mathcal{R}$ or $\mathcal{Z}$. It describes transmission in a population that has not been modified by immunity or control, and it counts every infectious bite that *would* cause an infection, even if the host is already infected.

$R_C$ - **The Adjusted Reproductive Number** is a scalar. In fact, $R_C$ is like $R_0$ in every way, except one – $R_C$ has been modified by malaria control. We think of $R_C$ as describing a family of numbers, of which $R_0$ is a special case where $C = 0$. The difference between $R_0$ and $R_C$ are related to the interpretation of $\mathcal{D}$ and $\mathcal{V}$. To compute $R_C$ , $D$ and $\mathcal{D}$ should be computed for a population that is lacking immunity, which may require changing some of the parameters describing transmission from strata that are immune.

$R_E$ - **The Endemic Reproductive Number** is a scalar. It describes transmission in a population that has been modified by immunity and control, but like $R_C$, it counts every infectious bite that *would* cause an infection, even if the host is already infected. The endemic number thus includes effect sizes of immunity on transmission, and $R_C/R_E$.

$R_e$ - **The Effective Reproductive Number** is a scalar. It describes transmission in a population that has been modified by immunity and control, but it does not count redundant infections – infections on a host that is already infected. The differences between $R_E$ and $R_e$ are entirely due to the fact that $R_E$ ignores the fact that some people might already be infected. For malaria, $R_e$ could be computed in various ways, depending on the model, but at the steady state of any model with endemic transmission, $R_e = 1$ by definition. We have not proposed any way of computing $R_e$ for this model.

- Local Adjusted Reproductive Numbers (or substituting $R_E$ for $R_C$, as appropriate, to get Local Endemic Reproductive Numbers):

  $\breve{R}_C$ - **Local Adjusted Reproductive Numbers**, $\left|\breve{R}_C\right| = p$ computed using a modified version of Macdonald's formula as if there was no movement of either humans or mosquitoes (Eq. 47).

  $\hat{R}_C$ - **Local Adjusted Reproductive Numbers**, $\left|\hat{R}_C\right| = p$, the number of infections arising from a patch starting from a single person infected in a each patch (Eq. 48).

  $\tilde{R}_C$ - **Local Adjusted Reproductive Numbers**, $\left|\hat{R}_C\right| = p$, the number of infections arising from a patch starting from a single mosquito infected in a each patch (Eq. 49).

#### Abbreviations

dEIR - daily **Entomological Inoculation Rate (EIR)**, Biting, $|\text{dEIR}| = n$. In this model, the true EIR among residents is $\beta \cdot fqvZ$ and for the visitors it is $(1 - v)fqZ$. (see Eq. 9).

dFoI - daily **Force of Infection (dFoI)**; $|h| = n$. The FoI is defined as the hazard rate for infections. Here, the dFoI is the sum of the local FoI, $h$ and the travel FoI, $\delta$.

dHBR - daily **Human Biting Rate**, $|\text{dHBR}| = n$. The human biting rate is defined as the expected number of bites by vector mosquitoes, per person, per day. In this model, the true HBR among residents from a single species is $\beta \cdot fqvM$ and for the visitors it is $(1 - v)fqM$.

dLHBR - daily **Local Human Biting Rate**, $|\text{dLHBR}| = p$. The expected number of bites in each patch, per person, per day, is $fqM/W$. It is computed for someone with a biting weight of one ($w_f = 1$).

dLEIR - daily **Local Entomological Inoculation Rate**, $|\text{dLEIR}| = p$. The expected number of bites in each patch, per person, per day, is $fqZ/W$. It is computed for someone with a biting weight of one ($w_f = 1$).

SR, $z$ - **Sporozoite Rate**, z; $|z| = p$. The fraction of mosquitoes in each patch with sporozoites in salivary glands and presumed infectious (Eq. 8).

### Alphabetical

#### Caligraphic Script, $\mathcal{A} - \mathcal{Z}$

$\mathcal{B}$ - A moniker for **The Blood Feeding Model**

$\mathcal{D}$ - **The Human Transmitting Capacity Distribution Matrix**, $|\mathcal{D}| = p \times p$. $\mathcal{D}$ computes a measure of total HTC jointly spent among the patches, which describes how parasites are dispersed among patches by the human population strata (Eq. 40).

$\mathcal{G}$ - **The next generation matrix**, $|\mathcal{G}| = (n+p) \times (n+p)$, computed as a "type reproductive number." Note that most of the analysis is done with the diagonal blocks, $\mathcal{R}$ and $\mathcal{Z}$, of the parasite's next generation matrix, $\mathcal{G}^2$.

$\mathcal{J}$ - **Residency Membership Matrix**, $|\mathcal{J}| = p \times n$. A matrix where the $i, j^{th}$ element is 1 if the $j^{th}$ population stratum resides in the $i^{th}$ patch. For example, $\mathcal{J} \cdot H = P$.

$\mathcal{K}$ - **Mosquito dispersal matrix**, $|\mathcal{K}| = p \times p$. The $i, j^{th}$ element of $\mathcal{K}$ is the fraction mosquitoes leaving the $i^{th}$ patch who end up in the $j^{th}$ patch.

$$\mathcal{K} = \left[ \begin{array}{ccccc} j=1 & j=2 & j=3 & \dots & j=p \\ \boxed{\begin{array}{c} 0 \\ \mathcal{K}_{2,1} \\ \mathcal{K}_{3,1} \\ \vdots \\ \mathcal{K}_{p,1} \end{array}} & \boxed{\begin{array}{c} \mathcal{K}_{1,2} \\ 0 \\ \mathcal{K}_{3,2} \\ \vdots \\ \mathcal{K}_{p,2} \end{array}} & \boxed{\begin{array}{c} \mathcal{K}_{1,3} \\ \mathcal{K}_{3,2} \\ 0 \\ \vdots \\ \mathcal{K}_{p,3} \end{array}} & \boxed{\begin{array}{c} \dots \\ \dots \\ \dots \\ \ddots \\ \dots \end{array}} & \boxed{\begin{array}{c} \mathcal{K}_{1,p} \\ \mathcal{K}_{2,p} \\ \mathcal{K}_{3,p} \\ \vdots \\ 0 \end{array}} \end{array} \right] \tag{1}$$

By definition, $\text{diag}(\mathcal{K}) = 0$ and $\sum_j \mathcal{K}_{i,j} \leq 1$. Columns that do not sum to one have a net loss of mosquitoes associated with migration from the $j^{th}$ patch.

$\mathcal{L}$ - **Immature Mosquito Module State Space**. A generic moniker for the state space defined for the immature mosquito component. The dynamics are given by a system of equations denoted $d\mathcal{L}/dt$.

$\mathcal{M}$ - **Adult Mosquito Module State Space**. A generic moniker for the state space defined for the adult mosquito component. The dynamics are given by a system of equations denoted $d\mathcal{M}/dt$.

$\mathcal{N}$ - **Aquatic Habitat Membership Matrix**, $|\mathcal{N}| = p \times l$. A matrix where the $i, j^{th}$ element is 1 if the $j^{th}$ aquatic habitat is found in the $i^{th}$ patch: $\mathcal{N} \cdot 1$ is the number of habitats found in each patch.

$\mathcal{R}$ - **The parasite's next generation matrix for human strata**, $|\mathcal{R}| = n \times n$. It is the upper on-diagonal block of $\mathcal{G}^2$, which describes transmission among strata in the parasite's next generation (Eq. 45).

$\mathcal{U}$ - **Egg Dispersal Matrix**, $|\mathcal{U}(\mathcal{N}, w_\nu)| = l \times p$. A matrix, derived from $\mathcal{N}$ and $w_\nu$ describing how eggs laid by adults in a patch are allocated among the habitats (Eq. 14).

$\mathcal{V}$ - **Vectorial Capacity Matrix**, $|\mathcal{V}| = p \times p$. A matrix that describes a patch-specific analogue of vectorial capacity. It describes the number of infective bites eventually arising in every other patch, on average, from all the mosquitoes blood feeding on a single person, on a single day, assuming all those mosquitoes became infected upon blood feeding (Eq. 39).

$\mathcal{X}$ - **Human Epidemiology State Space**. A generic moniker for a state space defining for parasite infections and immunity in humans. The dynamics are given by a system of equations denoted $d\mathcal{X}/dt$.

$\mathcal{Y}$ - **Mosquito Infection State Space**. A generic moniker for the state space defined for the component describing parasite infections of adult mosquitoes. The dynamics are given by a system of equations denoted $d\mathcal{Y}/dt$.

$\mathcal{Z}$ - **The parasite's next generation matrix for patches**, $|\mathcal{Z}| = p \times p$. It is the lower on-diagonal block of $\mathcal{G}^2$, which describes transmission among strata in the parasite's next generation among patches (Eq. 45).

#### Upper Case Roman, $A - Z$

$A$ - **Ambient human population density**, $|A| = p$. The density of humans present in a patch at a point in time (Eq. 2)

$A_r$ - **Ambient resident human population density**, $|A_r = (J \odot \Theta) \cdot H| = p$ describing the density of humans in each patch who reside in each patch, used as a diagnostic statistic for the time - spent matrix.

$A_n$ - **Ambient non-resident, non-visitor human population density**, $|A - A_r| = p$ describing the density of humans from the spatial domain who do not reside in the patch, used as a diagnostic statistic for the time - spent matrix.

$B$ - **Blood Host Availability (All Vertebrate Hosts)**, $|B| = p$. A measure of the of vertebrate hosts for blood feeding. It is used to compute the blood feeding rate, the human blood feeding fraction, and the emigration rate: $B = W + W_\delta + O^\zeta$.

$D$ - **Human Transmitting Capacity**, $|D| = n$. The HTC is interpreted as an equivalent number of days (hence $D$) a person spends fully infectious [1]. It is computed by adding up days spent partially infections as a part of a day. To compute any spatial metrics involving reproductive success, $D$ must be defined from the model $\mathcal{X}$.

$E$ - **dEIR**, described in the Abbreviations section above.

$F$ - **Functional Response**

$F_f$ - **Blood feeding functional response**, a function that accepts a vector describing total host availability, $W$, and returns a vector of blood feeding rates, $f$. The function has a maximum rate $f_x$ and a shape parameter $s_f$: See Eq. 5.

$F_\nu$ - **Egg laying functional response**, a function. $F_\nu$ accepts a vector describing habitat availability, $Q$, and returns a vector of per-capita egg laying rates, $\nu$. The function has a maximum rate $\nu_x$ and a shape parameter $s_\nu$. See Eq. 12.

$F_\sigma$ - **Emigration functional response**. A function that accepts vectors describing the availability of hosts, $W$, habitats, $Q$, and sugar sources, $S$ and returns a vector of emigration rates, $\sigma$. It is assumed that mosquitoes move in search of resources, so the more available, the less the mosquito moves, but it will move in search of any one of three resources, so the rate is a sum of three terms, each with a maximum, $\sigma_*$ and a shape parameter, $s_*$. See Eq. 19.

$F_h$ - **Exposure functional response**. A function describing the dFoI as a function of the dEIR [2]. For the model defined here, $h = bE$.

$G$ - **Density of Gravid Mosquitoes** $|G| = p$. A vector of variables describing the density of gravid mosquitoes. In this model, only gravid mosquitoes can lay eggs.

$H$ - **Human strata population density**, $|H| = n$. The resident population, $P$, is segmented into a set of population strata to reduce heterogeneity and improve the accuracy of the models.

$I$ - an **Identity Matrix**. The dimensions of an identity matrix depend on context.

$L$ - **Total immature mosquito population density**, $|L| = l$. A vector of variables describing lumped immature mosquito population density. The dynamics are described by Eq. 16.

$M$ - **Mosquito population density**, $|M| = p$. A vector of variables describing adult, female mosquito population density. The dynamics are defined by Eq. 21.

$O$ - **Other vertebrate host availability**, $|O| = p$. $O$ is a measure of the weighted, ambient population density of non - human vertebrate hosts term that is directly comparable to human availability, $W$.

$P$ - **The census population density**, $|P| = p$. The number of people who live in a patch, called residents. Some patches might have zero residents.

$Q$ - **Aquatic habitat availability**, $|Q| = p$. The availability of aquatic habitats in each patch is the patch-wise sum of the habitat search weights, $w_f$. It is used to compute egg laying rates, $\nu$, egg laying fractions, $\eta$, and mosquito emigration rates, $\sigma$. See Eq. 11.

$S$ - **Sugar Availability**, $|S| = p$. The availability of sugar affects mosquito movement. It is passed to the model as a vector of functions describing sugar availability over time.

$W$ - **Human availability**, $|W| = p$. A measure of availability for blood feeding. It is a weighted measure of ambient population density, where the weights could take into account factors such as body size, ITN usage, or habitual use of personal protection. It is used in the computation of $\beta$, mosquito movement, including both the rate of emigration $\sigma$ and the dispersal kernel, $K$.

$W_\delta$ - **Availability of Visitors**, $|W_\delta| = p$. Availability of visitors from outside the spatial domain.

$X$ - **Infective Density of Infectious Humans**, $|X| = n$. A dynamical quantity describing the effective infectious density of each stratum. This relationship must be defined in relation to the state space, $\mathcal{X}$. Here, $X = xH$.

$Y$ - **Density of infected mosquitoes**, $|Y| = p$. A variable describing the density of infected mosquitoes, defined by Eq. 23.

$Z$ - **Infective Density of Infectious Mosquitoes**, $|Z| = p$. A dynamical quantitity. $Z$ is a variable describing the density of blood feeding, infective mosquitoes, defined by Eq. 25.

#### Lower Case Roman, $a - z$

$b$ - **Transmission efficiency**, either $b$ is a scalar or $|b| = n$. The fraction of infective bites that cause an infection.

$c$ - **Infectiousness**, $|c| = n$. either $b$ is a scalar or $|b| = n$. The fraction of infective bites that cause an infection.

$c_1$ - is the infectiousness of a person with a patent infection.

$c_2$ - is the infectiousness of a person with a sub - patent infection.

$d$ - **Time of day**. $d = t - \text{floor}(t)$

$f$ - **Mosquito blood feeding rate**, $|f| = p$. In each patch, the number of blood meals, per mosquito, per day (Eq. 5).

$g$ - **Mosquito mortality rates**, $|f| = p$. In each patch, the death rate of mosquitoes, per mosquito, per day.

$h$ - **Local Force of Infection**, $|h| = n$. A term describing stratum specific hazard rate for local infection, the number of infections, per person, per day: The local FoI is computed from the dEIR $h = f_h(E)$. For the model family defined in this paper, $h = bE$.

$k$ - **Stratum infectiousness**, $|k| = n$. A term describing the proportion of mosquitoes that become infected after blood feeding on a mosquito from each stratum (Eq. 30).

$k_\delta$ - **Visitor's infectiousness**, $|k_\delta| = p$. The fraction of mosquitoes that become infected after blood feeding on a visitor.

$l$ - **The number of aquatic habitats**, $|l| = 1$.

$m_1$ - **Mean true multiplicity of infection (MoI)**, $|m_1| = n$. A vector of variables describing the mean true MoI in each population stratum. The dynamics of $m$ are defined by Eq. 26. In this model, true prevalence is $x_1 = 1 - e^{-m_1}$.

$m_2$ - **Mean apparent multiplicity of infection (MoI)**, $|m_2| = n$. A vector of variables describing the mean apparent MoI in each stratum. The dynamics of $m_2$ are defined by Eq. 28. In this model, apparent prevalence is $x_2 = 1 - e^{-m_2}$

$n$ - **The number of human population strata**, $|n| = 1$

$p$ - **The number of patches**, $|p| = 1$

$q$ - **The human blood feeding fraction** or **human fraction**, $|q| = p$. The fraction of blood meals in which the mosquito fed on a human (Eq. 6).

$r_1$ - **Clearance rate**, $|r_1| = n$. The duration of a infection is $1/r_1$ days, on average.

$r_2$ - **Apparent Clearance rate**, $|r_2| = n$. The duration of an apparent infection is $1/r_2$ days, on average.

$t$ - **Time**.

$w$ - **Search weights**

- $w_f$ - Blood feeding search weights, a vector of length $n$, are used to compute the availability of humans in each strata (See Eq. 3). The search weights can be used to modify exposure to model usage of insecticide treated nets, differences in exposure related to body size or risky activities, use of personal protection devices, or other factors that modify blood feeding or exposure.
- $w_\nu$ - Egg laying search weights for aquatic habitats, a vector of length $l$. These search weights are used to compute the total availability of habitats, $Q$ (see Eq. 11). By using search weights to compute availability, it is possible to modify a baseline to consider various modes of control. Two possibilities are adding devices, such as oviposition traps, or fouling the water in a habitat.

$x$ - **Infectiousness**, $|x| = n$. The probability a mosquito blood feeding on a human in each stratum becomes infected.

$x_1$ - **True prevalence**, $|x_1| = n$. A term describing the true prevalence of *P. falciparum* infection in each population stratum (Eq. 27)

$x_2$ - **Apparent prevalence**, $|x_2| = n$. A term describing the apparent prevalence of *P. falciparum* infection in each population stratum (Eq. 29).

$x_\delta$ - **Infectiousness, visitors**, $|x| = n$. The probability a mosquito blood feeding on a human visiting from outside the spatial domain becomes infected.

#### Upper Case Greek, $A - \Omega$

$\Gamma$ - **Patch Egg Laying Rate**, $|\Gamma| = p$. The total egg laying rate is $\Gamma = \chi \nu G$.

$\Theta$ - **Time Spent Matrix**, $|\Theta| = p \times n$. The $j^{th}$ column of $\Theta$ describes time spent by humans among patches.

$$\Theta = \left[ \begin{array}{ccccc} j=1 & j=2 & j=3 & \cdots & j=p \\ \boxed{\begin{array}{c} \Theta_{1,1} \\ \Theta_{2,1} \\ \Theta_{3,1} \\ \vdots \\ \Theta_{p,1} \end{array}} & \boxed{\begin{array}{c} \Theta_{1,2} \\ \Theta_{2,2} \\ \Theta_{3,2} \\ \vdots \\ \Theta_{p,2} \end{array}} & \boxed{\begin{array}{c} \Theta_{1,3} \\ \Theta_{3,2} \\ \Theta_{3,3} \\ \vdots \\ \Theta_{p,3} \end{array}} & \boxed{\begin{array}{c} \cdots \\ \cdots \\ \cdots \\ \ddots \\ \cdots \end{array}} & \boxed{\begin{array}{c} \Theta_{1,p} \\ \Theta_{2,p} \\ \Theta_{3,p} \\ \vdots \\ \Theta_{p,p} \end{array}} \end{array} \right] \tag{2}$$

$\Xi$ - **A Mosquito tracking matrix**, $|\Xi| = p \times p$. The mosquito tracking matrix describes the location of cohorts of mosquitoes over time (Eq.!37).

$\Upsilon$ - **Survival and dispersal through the EIP**, $|\Upsilon_\tau| = p \times p$. $\Upsilon_\tau(t)$ describes mosquito dispersal and survival from $t - \tau$ to $t$ (Eq. 24). If $\Omega$ is constant then

$$\Upsilon_\tau(t) = e^{-\Omega\tau}.$$

If $\Omega(t)$ is reasonably smooth over the period, a reasonable approximation could be $\Upsilon_\tau(t) = e^{-\Omega(t-\tau/2)}$.

$\Psi$ - **Time at Risk Matrix**, $|\Psi| = p \times n$. The time at risk matrix modifies time spent by the mosquito circadian pattern,

$$\Psi(t) = \text{diag}\left(\xi(t)\right) \cdot \Theta(t).$$

$\Lambda$ - **Patch Emergence rate**, $|\Lambda| = p$. A dynamical quantity. The number of adult, female mosquitoes emerging, per patch, per day, $\Lambda = \mathcal{N}\alpha$

$\Omega$ - **Mosquito demography matrix**, $|\Omega| = p \times p$. A matrix describing mosquito mortality and movement rates among the patches. See Eq. 20.

#### Lower Case Greek, $\alpha - \omega$

$\alpha$ - **Habitat emergence rate**, $|\alpha| = l$. The number of adult, female mosquitoes emerging, per habitat, per day. A model for aquatic mosquito populations must compute $\alpha$ from the aquatic state space $\mathcal{L}$.

$\beta$ - **Biting distribution matrix**, $|\beta| = n \times p$. The matrix $\beta$ describes the proportion of each bite occurring in each patch that is received by an individual in each stratum, defined by Eq. 7. For example, (setting $\upsilon = 1$), to compute the EIR for all the strata, we write $\beta \cdot fqZ$. An important constraint is that the columns of

$$\text{diag}(H) \cdot \beta$$

sum up to 1; $\beta$ allocates bites to *individuals* in each stratum. To compute the FoI for mosquitoes (again, setting $\upsilon = 1$), we write it in several different ways:

$$fq\kappa = fq\beta^T \cdot X = \text{diag}\left(\frac{fq}{W}\right) \cdot \Psi^T \cdot w_f xH = fq\frac{\Psi^T \cdot w_f xH}{\Psi^T \cdot w_f H}.$$

$\gamma$ - **Net visitor infectivity**, $|\gamma| = p$. In each patch, the fraction of infected mosquitoes that were infected by visitors.

$\delta$ - **Travel FoI**, $|\delta| = n$. In each stratum, the net rate of infection from travel outside the spatial domain.

$\zeta$ - **Host preference**. Usually, $|\zeta| = 1$, but with patch - specific scaling, $|\zeta| = p$. A shape parameter describing a power - law scaling relationship of total availability to non - human host availability (see $W$)

$\eta$ - **Egg Distribution Rates**, $|\eta| = l$. The egg laying rate in each aquatic habitat (Eq. 15).

$\theta$ - **Density dependent mortality for larvae**, $|\theta| = l$. Total mortality rate for immature mosquitoes in each patch is $\phi + \theta L$.

$\kappa$ - **Net infectiousness**, $|\kappa| = p$. The proportion of mosquitoes in each patch who become infected after blood feeding on a human (Eq. 10).

$\sigma$ - **Emigration rate**, $|\sigma| = p$. The net immigration rate of mosquitoes from each patch is assumed to be a function of unsuccessful search for resources. See Eq. 19.

$\nu$ - **The population egg laying rate**, $|\nu| = l$. The egg laying rate by the adult mosquito population in each patch, per day (Eq. 13). Eggs are distributed among the patches by availability (see $\eta$).

$\xi$ - **Circadian weight function**, $|\xi(d)| = 1$. The function describes the relative blood feeding activity rates of mosquitoes. It is constrained such that $\int_0^1 \xi(s)ds = 1$

$\tau(t)$ - **Extrinsic Incubation Period (EIP)**, $|\tau(t)| = 1$. The number of days required for an infected mosquito infected at time $t$ to become infective. We also define $\tau^{-1}(t)$, which is the time a mosquito becoming infective at time $t$ became infected.

$\phi$ - **Density independent mortality for larvae**, $|\theta| = l$. The death rate from all density - independent causes.

$\chi$ - A scalar. The number of eggs laid, per batch.

$\psi$ - **Maturation rate for immature mosquitoes**, $|\psi| = l$. The rate immature mosquitoes mature. If $\psi$ is constant then the time from egg to emergence is $1/\psi$.

$\upsilon$ - **Resident Fraction**. $|\upsilon| = p$. The fraction of biting that occurs on residents. See Eq. 4. With no malaria importation, we set $\upsilon = 1$.
