## Supplement 3 - Modular Notation for "Spatial Dynamics of Malaria Transmission"

To do modular computation, we need to develop a *modular notation* as a way of encoding a model. This vignette is designed to explain modular notation by constructing a model with five aquatic habitats ($l = 5$), three patches ($p = 3$), and four human population strata ($n = 4$). We call it `5-3-4`.

First, we present `5-3-4` using conventional notation (the **Conventional** tab/section), and then we rewrite it using modular notation (the **Modular** tab/section). The middle section (the **Transform** tab/section) walks through the differences step by step.

For a discussion of solving, see `ModularComputation.Rmd` or ModularComputation.html.

### Diagram

The model `5-3-4` is designed to illustrate some important features of the framework and notation. We assume that:

- the first three habitats are found in patch 1; the last two are in patch 2; patch 3 has no habitats.
- patch 1 has no residents; patches 2 and 3 are occupied, each with two different population strata;
- Transmission among patches is modeled using the concept of *time spent*, which is similar to the *visitation rates* that have been used in other models. While the strata have a residency, each stratum can spend some time in each habitat.

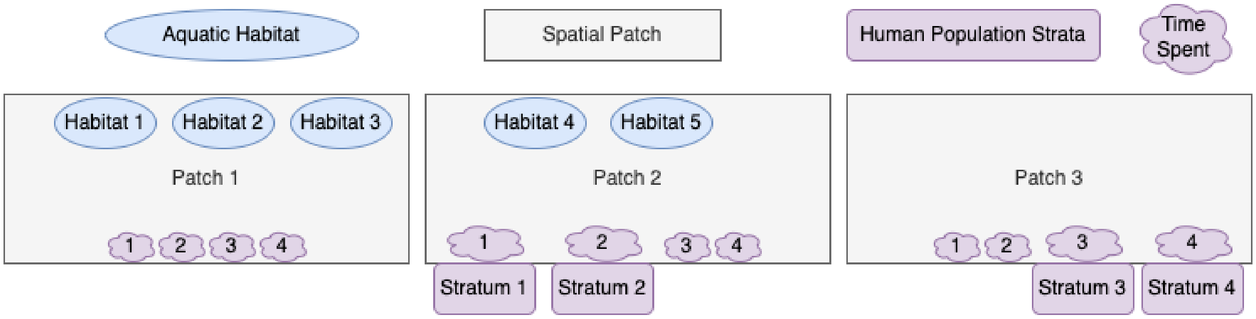


### Conventional

The model `5-3-4` is presented here in conventional notation as a set of four linked dynamical components:

#### Aquatic Dynamics

We let $L_{i,j}$ denote an aquatic population in the $j^{th}$ habitat in patch $i$. Similarly, we let $\psi_{i,j}$ denote the maturation rate, $\phi_{i,j}$ the density independent mortality rate, and $\theta_{i,j} L_{i,i}$ the density dependent increase in mortality rates in response to mean crowding. We let $\Gamma_i$ denote the daily egg laying rates in each patch, and we let $\xi_{i,j}$ be the fraction of eggs in the $i^{th}$ patch that are laid in the $j^{th}$ habitat.

The following system of equations describes mosquito aquatic dynamics in five habitats. The first three are found in one patch, and the last two are found in the second patch. The third patch has no aquatic habitats:

$$\begin{aligned}
dL_{1,1}/dt &= \xi_{1,1}\Gamma_1 - (\psi_{1,1} + \phi_{1,1} + \theta_{1,1}L_{1,1})L_{1,1} \\
dL_{1,2}/dt &= \xi_{1,2}\Gamma_1 - (\psi_{1,2} + \phi_{1,2} + \theta_{1,2}L_{1,2})L_{1,2} \\
dL_{1,3}/dt &= \xi_{1,3}\Gamma_1 - (\psi_{1,3} + \phi_{1,3} + \theta_{1,3}L_{1,3})L_{1,3} \\
dL_{2,1}/dt &= \xi_{2,1}\Gamma_2 - (\psi_{2,1} + \phi_{2,1} + \theta_{2,1}L_{2,1})L_{2,1} \\
dL_{2,2}/dt &= \xi_{2,2}\Gamma_2 - (\psi_{2,2} + \phi_{2,2} + \theta_{2,2}L_{2,2})L_{2,2}
\end{aligned} \tag{1}$$

We assume that $\alpha_{i,j} = \psi_{i,j}L_{i,j}/2$, and

**Adult Mosquito Dynamics**

We let $M_i$ denote adult mosquito density in patches $i = 1, 2, 3$. We let $g_i$ denote per-capita mortality, $\sigma_i$ denote the emigration rate, and $k_{i,j}$ the fraction of emigrating mosquitoes that move from $i$ to $j$. Recruitment from aquatic habitats is the sum of emergence rates, so we write:

$$\begin{aligned}
\frac{dM_1}{dt} &= \sum_j \alpha_{1,j} - g_1M_1 - \sigma_1M_1 + k_{2,1}\sigma_2M_2 + k_{3,1}\sigma_3M_3 \\
\frac{dM_2}{dt} &= \sum_j \alpha_{2,j} - g_2M_2 - \sigma_2M_2 + k_{1,2}\sigma_1M_1 + k_{3,2}\sigma_3M_3 \\
\frac{dM_3}{dt} &= \sum_j \alpha_{3,j} - g_3M_3 - \sigma_3M_3 + k_{1,3}\sigma_1M_1 + k_{2,3}\sigma_2M_2
\end{aligned} \tag{2}$$

Noting that $\sum_j \alpha_{3,j} = 0$ because there are no habitats in patch 3, so it is actually reduntant.

We have developed a model for gravid mosquitoes to model egg laying rates. We let $G_i$ denote the density of gravid / egg laying mosquitoes in in patches $1, 2, 3$.

We let $\nu_i$ denote the per-capita egg-laying rate, and $f_i$ denote the blood feeding rate.

$$\begin{aligned}
\frac{dG_1}{dt} &= f_1(M_1 - G_1) - \nu_1G_1 - g_1G_1 - \sigma_1G_1 + k_{2,1}\sigma_2G_2 + k_{3,1}\sigma_3G_3 \\
\frac{dG_2}{dt} &= f_2(M_2 - G_2) - \nu_2G_2 - g_2G_2 - \sigma_2G_2 + k_{1,2}\sigma_1G_1 + k_{3,2}\sigma_3G_3 \\
\frac{dG_3}{dt} &= f_3(M_3 - G_3) - \nu_3G_3 - g_3G_3 - \sigma_3G_3 + k_{1,3}\sigma_1G_1 + k_{2,3}\sigma_2G_2
\end{aligned} \tag{3}$$

To connect egg laying by adults to eggs deposited, we let $\Gamma_i = \chi\nu_iG_i$, where $\chi$ is the number of eggs laid, per batch. We would write $\Gamma_3 = \chi\nu_3G_3$ but if there are really no habitats, then mosquitoes must leave a patch to blood feed, so $\Gamma_3 = 0$.

**Parasite Infection Dynamics in Mosquitoes**

We let $Y_i$ denote the density of infected mosquitoes in patches $1, 2, 3$.

To model infection dynamics, we let $\kappa_i$ denote the probability a mosquito would become infected after blood feeding on a human. In a moment, we will describe how this is computed, but for now, we can write:

$$\begin{aligned}
\frac{dY_1}{dt} &= f_1q_1\kappa_1(M_1 - Y_1) - g_1Y_1 - \sigma_1Y_1 + k_{2,1}\sigma_2Y_2 + k_{3,1}\sigma_3Y_3 \\
\frac{dY_2}{dt} &= f_2q_2\kappa_2(M_2 - Y_2) - g_2Y_2 - \sigma_2Y_2 + k_{1,2}\sigma_1Y_1 + k_{3,2}\sigma_3Y_3 \\
\frac{dY_3}{dt} &= f_3q_3\kappa_3(M_3 - Y_3) - g_3Y_3 - \sigma_3Y_3 + k_{1,3}\sigma_1Y_1 + k_{2,3}\sigma_2Y_2
\end{aligned} \tag{4}$$

Here we want to model adult mosquito density as a term that accounts for a delay, but without implementing a delay differential equation. Instead we assume that $Z$ is a function of $Y$, but it discounts for surviving and dispersing through the EIP, which lasts $\tau$ days (by assumption). Without movement, we would get that $Z_i = e^{-g_i\tau}Y_i$, but with movement, we must develop a matrix that computes survival and dispersal through the EIP. To do so, we let $\mathcal{K}$ denote the dispersal matrix (using the terms from above) describing where emigrating mosquitoes leave.

$$\mathcal{K} = \left[\begin{array}{ccc} 0 & k_{2,1} & k_{3,1} \\ k_{1,2} & 0 & k_{3,2} \\ k_{1,3} & k_{2,3} & 0 \end{array}\right]$$

Now, we can formulate a single matrix that describes survival and dispersal, denoted $\Omega$:

$$\Omega = \text{diag}(g) + (I - \mathcal{K}) \cdot \text{diag}(\sigma)$$

where $e^{-\Omega\tau}$ describes survival and dispersal of a cohort of mosquitoes through the EIP. Matrix exponentiation is computed in `R` using the function `expm`. We let $Z = e^{-\Omega\tau}Y$ denote the density of infective mosquitoes.

**Blood Feeding and Transmission**

We have already developed a model of mosquito mobility. Unlike mosquitoes, humans have a home, so we follow a different set of rules using a matrix that describes time spent, $\Theta$. The rows of $\Theta$ represent patches and the columns of $\Theta$ represent human population strata. We let $\Theta_{i,j}$ denote the fraction of a day spent by the $j^{th}$ population stratum in the $i^{th}$ patch. Each column of $\Theta$ (looking across the index $i$ for a fixed $j$) represents time spent by each one of the strata in each one of the populations. There are four strata and three patches, so:

$$\Theta = \left[\begin{array}{cccc} \Theta_{1,1} & \Theta_{1,2} & \Theta_{1,3} & \Theta_{1,4} \\ \Theta_{2,1} & \Theta_{2,2} & \Theta_{2,3} & \Theta_{2,4} \\ \Theta_{3,1} & \Theta_{3,2} & \Theta_{3,3} & \Theta_{3,4} \end{array}\right]$$

As we have defined this model, there are four strata and three patches. The first two strata are found in patch 2, and the second two strata are found in patch 3. This is a somewhat unconventional way of modeling humans – most models for malaria spatial dynamics have a one patch, one stratum rule – but if we do not allow for humans in patches to be segmented, then the structure of the model enforces a rule that humans in patches are homogeneous. In fact, those populations could differ in many ways that are important for transmission. We must now develop a notation for dealing with the human strata.

One way to build the model is to index population density by their residency, patch 2 first stratum would have index $2, 1$, and we now have a population vector:

$$H = \left[\begin{array}{c} H_{2,1} \\ H_{2,2} \\ H_{3,1} \\ H_{3,2} \end{array}\right].$$

The challenge is to keep this notation while we describe where these humans spend their time. The information about residency ends up being used in setting the parameter values in $\Theta$, but now the $j^{th}$ column in $\Theta$ would map onto a population with two indices. In this new model, residency is somewhat irrelevant for modeling transmission because all the information we need to model transmission is stored in the time spent matrix. The notation easier if we use a single index, so we let $H_i$ be population density of each stratum.

$$H = \left[\begin{array}{c} H_1 \\ H_2 \\ H_3 \\ H_4 \end{array}\right].$$

Here, we assume humans are all equally likely to be bitten, so the ambient density of humans, which we will call availability, is

$$W_i = \sum_j \Theta_{i,j} H_j.$$

In this model, availability is equal to ambient density because all humans are bitten at the same rate. Later, we will introduce biting weights so that the strata could be exposed at different rates, reflecting differences in exposure due to age, net use, or other factors.

We assume that $q_i$ denotes the fraction of bites taken on a human, so $f_i q_i Z_i$ is the net daily human blood feeding rate, and there are

$$f_i q_i \frac{Z_i}{W_i}$$

bites per person in that patch. We could call $E_i$ the daily EIR, but this is not what a person in a stratum would experience. Instead, a person in the $j^{th}$ stratum spends some time in every patch, so for the $j^{th}$ stratum, the rate of exposure is:

$$E_j = \sum_{i=1}^{3} \Theta_{i,j} f_i q_i \frac{Z_i}{W_i}$$

Note that the index on $E$ describes a population stratum, not a patch.

We also need to define $\kappa_i$, the probability a mosquito in the $i^{th}$ patch would become infected after biting a human. To do so, we let $x_j$ denote the probability a mosquito would become infected after biting a human in the $j^{th}$ stratum. Now, we are summing in a patch across stratum and we let:

$$\kappa_i = \frac{\sum_{j=1}^{4} \Theta_{i,j} x_j H_j}{\sum_{j=1}^{4} \Theta_{i,j} H_j}.$$

**Infection Dynamics in Humans**

We let $X_i$ denote the density of infected and infectious humans, and $r$ the rate that infections clear. We let $b$ denote the fraction of infective bites that cause an infection. Now, a person's risk is computed by adding the product of their time at risk and the daily EIR across all three patches:

$$\begin{aligned}
\frac{dX_1}{dt} &= b \sum_{i=1}^{3} \Theta_{i,1} f_i q_i \frac{Z_i}{W_i} (H_1 - X_1) - rX_1 \\
\frac{dX_2}{dt} &= b \sum_{i=1}^{3} \Theta_{i,2} f_i q_i \frac{Z_i}{W_i} (H_2 - X_2) - rX_2 \\
\frac{dX_3}{dt} &= b \sum_{i=1}^{3} \Theta_{i,3} f_i q_i \frac{Z_i}{W_i} (H_3 - X_3) - rX_3 \\
\frac{dX_4}{dt} &= b \sum_{i=1}^{3} \Theta_{i,4} f_i q_i \frac{Z_i}{W_i} (H_4 - X_4) - rX_4
\end{aligned} \tag{5}$$

Or equivalently, using $E_i$:

$$\begin{aligned}
\frac{dX_1}{dt} &= bE_1(H_1 - X_1) - rX_1 \\
\frac{dX_2}{dt} &= bE_2(H_2 - X_2) - rX_2 \\
\frac{dX_3}{dt} &= bE_3(H_3 - X_3) - rX_3 \\
\frac{dX_4}{dt} &= bE_4(H_4 - X_4) - rX_4
\end{aligned} \tag{6}$$

Finally, to complete the model, we need to compute $\kappa_i$ for each patch, which is used by the model describing parasite infection dynamics in mosquitoes.

We let $x_j = c_j X_j / H_j$ denote the fraction of bites on an individual from the $j^{th}$ stratum that would infect the mosquito. Since $x_j H_j = c_j X_j$, we can rewrite $\kappa_j$ using :

$$\begin{aligned}
\kappa_1 &= \frac{\sum_j \Theta_{1,j} c_j X_j}{\sum_j \Theta_{1,j} H_j} \\
\kappa_2 &= \frac{\sum_j \Theta_{2,j} c_j X_j}{\sum_j \Theta_{2,j} H_j} \\
\kappa_3 &= \frac{\sum_j \Theta_{3,j} c_j X_j}{\sum_j \Theta_{3,j} H_j}
\end{aligned} \tag{7}$$

These $\kappa_i$ are used in the equations $dY_i/dt$ above.

We have now fully specified the model that we call 5-3-4 using traditional notation.

### Transform

In this section, we rewrite the model `5-3-4` with modular notation. To see the **full** model in *modular* notation, which directly parallels the *conventional* model, skip ahead to the next tab.

Here, we go step by step through each component, and we describe how the new notation facilitates modular construction of models.

#### Aquatic Dynamics ($\mathcal{L}$)

Here we describe how to translate the aquatic mosquito population model from conventional notation into modular notation, which facilitates modular computation.

**Reindex** In the modular notation, we re-index from double indices to a single index. In the triplet below, the first column is the old set of indices, which maps to ($\mapsto$) the middle column is the new set of indices, and the last column – called the membership vector – is the patch each aquatic habitat is found in ($\in$); the [old index] $\mapsto$ [new index] $\in$ [patch index]

$$\begin{bmatrix} 1,1 \\ 1,2 \\ 1,3 \\ 2,1 \\ 2,2 \end{bmatrix} \mapsto \begin{bmatrix} 1 \\ 2 \\ 3 \\ 4 \\ 5 \end{bmatrix} \in \begin{bmatrix} 1 \\ 1 \\ 1 \\ 2 \\ 2 \end{bmatrix} \tag{8}$$

This membership vector is, in fact, the most compact way of representing patch membership. For purposes of *encoding* a model, we need only store the membership vector; the same file could store parameters, or any other properties of the habitat.

Since all variables and parameters in modular notation have a single index; the new modular index no longer conveys information about location. Instead, for computational purposes, information about location is stored in the membership matrix, $\mathcal{N}$. In modular notation, we let the $i^{th}$ element is the new index, and the $j^{th}$ element the corresponding entry in the membership vector, and the $i, j^{th}$ element of the membership matrix is 1.

$$\mathcal{N} = \begin{bmatrix} 1 & 1 & 1 & 0 & 0 \\ 0 & 0 & 0 & 1 & 1 \\ 0 & 0 & 0 & 0 & 0 \end{bmatrix} \tag{9}$$

Using this, we re-index all the parameters.

We introduce a new term $\eta$ that describes egg deposition rates in the habitats, where

$$\eta_i = \xi_{i,j}\Gamma_i$$

denote the egg laying rate if any eggs are laid, and $\eta_i = 0$ otherwise. The dynamics are now described by the system of $i = 1, 2, ...5$ differential equations:

$$\tfrac{dL_i}{dt} = \eta_i - (\psi_i + \phi_i + \theta_i L_i) L_i \tag{10}$$

and we let

$$\alpha_i = \psi_i L_i / 2 \tag{11}$$

denote the emergence rate of adult, female mosquitoes.

**Vectorize** All our variables and parameters are defined as vectors. For example, we let $L$ denote the column vector:

$$L = \begin{bmatrix} L_1 \\ L_2 \\ L_3 \\ L_4 \\ L_5 \end{bmatrix} \tag{12}$$

In the same way, we represent $\psi$, $\phi$, $\theta$, and $\alpha$ as vectors.

Similarly, we let $\Gamma$ be a vector describing egg laying rates. We want to compute egg laying, so we redefine the vector $\xi$:

$$\xi = \begin{bmatrix} \xi_{1,1} \\ \xi_{1,2} \\ \xi_{1,3} \\ \xi_{2,1} \\ \xi_{2,2} \end{bmatrix} \tag{13}$$

Note that we have constrain parameters such that every egg is laid in a habitat,

$$\sum_j \xi_{i,j} = 1$$

which is equivalent to:

$$\mathcal{N}^T \cdot \xi = \text{sign}(\Gamma)$$

where sign returns a 1 if any eggs are laid, and 0 otherwise.

We let $\mathcal{U}$ denote the egg distribution matrix:

$$\mathcal{U} = \left(\mathcal{N} \cdot \text{diag}(\xi)\right)^T = \begin{bmatrix} \xi_{1,1} & 0 & 0 \\ \xi_{1,2} & 0 & 0 \\ \xi_{1,3} & 0 & 0 \\ 0 & \xi_{2,1} & 0 \\ 0 & \xi_{2,2} & 0 \end{bmatrix} \tag{14}$$

Finally, we let

$$\eta = \mathcal{U} \cdot \Gamma$$

describe egg deposition rates. As described above, the new parameter $\eta_i$ replaces $\xi_{i,j} \cdot \Gamma_i$, and we represent $\eta$ in vector form.

**Model** Egg laying rates, $\Gamma$, are computed by $\mathcal{M}$, passed to $\mathcal{L}$, and transformed into egg deposition rates, $\eta$:

$$\eta = \mathcal{U} \cdot \Gamma \tag{15}$$

The aquatic dynamics are described by the equations:

$$\frac{dL}{dt} = \eta - (\psi + \phi + \theta L)L \tag{16}$$

Emergence rates from the aquatic habitats are defined, transformed.

$$\begin{array}{ll} \alpha & = \frac{\psi L}{2} \\ \Lambda & = \mathcal{N} \cdot \alpha \end{array} \tag{17}$$

The vector $\Lambda$ is passed back to $\mathcal{M}$:

**Generic** If we take a birds-eye view of model building, we can describe model specification as an instance of a generic process:

- Define the model structure in relation to other components:
  - Define the habitat membership matrix, $\mathcal{N}$, that relates the structural elements of $\mathcal{M}$ ($p$ patches) and $\mathcal{L}$ ($l$ habitats)
  - Define model inputs. In this case, $\Gamma$ are the egg laying rates, computed by $\mathcal{M}$ and passed to $\mathcal{L}$.
  - We need to specify how eggs are distributed among the patches. We define a vector $\xi$, where
  $$\mathcal{N}^T \cdot \xi = 1.$$
  - We compute the egg-laying matrix, $\mathcal{U}$:
  $$\mathcal{U} = \mathcal{N}^T \cdot \text{diag}(\xi).$$
  - Egg laying rates (in patches) is transformed into egg deposition rates (in habitats) using the egg distribution matrix, $\mathcal{U}$:
  $$\eta = \mathcal{U} \cdot \Gamma$$
- Define the dynamics, $dL/dt$.
  - Define the state space. In this case, the model variables are fully defined by a column vector $L$ of length $l$.
  - Define model parameters. In this case, $\psi$ are per-capita maturation rates, and $\phi + \theta L$ per-capita mortality rates. All parameters are vectors of length $l$.
  - The dynamics are described by a system of ordinary differential equations:
  $$\frac{dL}{dt} = \eta - (\psi + \phi + \theta L)L \tag{18}$$
- Define the outputs of $\mathcal{L}$
  - We define a term describing the emergence of adults from habitats $\alpha$. In this case, $\alpha = \psi L/2$.
  - The net emergence rate of adult mosquitoes, per patch, which is computed by $\mathcal{L}$ and passed to $\mathcal{M}$, is
  $$\Lambda = \mathcal{N} \cdot \alpha.$$

**Adult Mosquitoes ($\mathcal{M}$ & $\mathcal{Y}$)**

Here we describe how to translate the mosquito ecology and parasite infection dynamics models from conventional notation into modular notation, which facilitates modular computation.

**Vectorize** There is no need to re-index $\mathcal{M}$, in part, because the other components are defined to interact with the patches. Most of the
work is done by defining $\mathcal{N}$ in the interface with $\mathcal{L}$, or in defining the mixing matrix $\beta$ and the computation of $\kappa_i$

To simplify the equations, once again, we introduce matrix notation. Let

$$M = \begin{bmatrix} M_1 \\ M_2 \\ M_3 \end{bmatrix}$$

and we also transform $G$ and $Y$ into column vectors. Similarly, we turn $f$, $\nu$, and $\kappa$ into column vectors.

We note that in defining the relationship between $Y$ and $Z$, we defined a demographic matrix $\Omega$. To recap, we let $g$ and $\sigma$ denote vectors describing survival in and emigration from the patches. The dispersal matrix, $\mathcal{K}$ is defined by

$$\mathcal{K} = \begin{bmatrix} 0 & k_{2,1} & k_{3,1} \\ k_{1,2} & 0 & k_{3,2} \\ k_{1,3} & k_{2,3} & 0 \end{bmatrix}$$

With these, we define

$$\Omega = \text{diag}(g) + (I - \mathcal{K}) \cdot \text{diag}(\sigma)$$

We can now write the equations describing adult mosquitoes, using $\Lambda$ as defined in $\mathcal{L}$, $\kappa$ as defined in $\mathcal{X}$ (below), and $\Omega$:

$$\begin{aligned} \frac{dM}{dt} &= \Lambda - \Omega \cdot M \\ \frac{dG}{dt} &= f(M - G) - \nu G - \Omega \cdot G \\ \frac{dY}{dt} &= fq\kappa(M - Y) - \Omega \cdot Y \end{aligned} \tag{19}$$

One core output of this dynamical component is egg laying

$$\Gamma = \chi \nu G \tag{20}$$

The other output is

$$Z = e^{-\Omega\tau} Y \tag{21}$$

**Generic** If we take a birds-eye view of model building, we can describe model specification as an instance of a generic process:

- Define the model structure
  - Define the patches.
  - Define model inputs from $\mathcal{L}$. In this case, $\Lambda$ are the emergence rates, computed by $\mathcal{L}$ and passed to $\mathcal{M}$.

  - Define model inputs from $\mathcal{X}$. In this case, $\kappa$ describes NI. It is computed by $\mathcal{B}$ and $\mathcal{X}$ and passed to $\mathcal{M}$.
- Define the dynamics, $dM/dt$.
  - Define the state space. In this case, the model variables are fully defined by column vectors of length $p$: adult mosquitoes, $M$; gravid mosquitoes $G$; and infected mosquitoes $Y$.
  - Define model parameters. In this case, $g$, $\sigma$, and the dispersal matrix $\mathcal{K}$, which are transformed into the demographic matrix $\Omega$:

$$\Omega = \text{diag}(g) + (I - \mathcal{K}) \cdot \text{diag}(\sigma)$$

  - We also define the parameter vectors $\nu$, $f$, $q$ and a scalar parameter $\chi$
  - The dynamics are described by a system of ordinary differential equations:

$$\begin{aligned} \frac{dM}{dt} &= \Lambda - \Omega \cdot M \\ \frac{dG}{dt} &= f(M - G) - \nu G - \Omega \cdot G \\ \frac{dY}{dt} &= fq\kappa(M - Y) - \Omega \cdot Y \end{aligned} \tag{22}$$

- Define the outputs of $\mathcal{M}$
  - We define egg laying rates as $\Gamma = \chi\nu G$
  - We define the density of blood feeding mosquitoes, $Z = e^{-\Omega\tau}Y$

**Human Infections ($\mathcal{X}$)**

**Blood Feeding ($\mathcal{B}$)** Because we resisted using double indices, we no longer have to worry about re-indexing. We find it useful, however, to store information about residency in a membership matrix:

$$\mathcal{J} = \begin{bmatrix} 0 & 0 & 0 & 0 \\ 1 & 1 & 0 & 0 \\ 0 & 0 & 1 & 1 \end{bmatrix}$$

The blood feeding model transforms a time spent matrix, $\Theta$, and a vector $H$ describing densities into a vector $W$ (of length $p$) describing availability of humans in patches:

$$W = \Theta \cdot H$$

Note that if we take $\mathcal{J} \cdot \mathcal{H}$, then we get the total density of residents, or the census population. If we take $(\mathcal{J} \odot \times) \cdot H$, where $\odot$ is the elementwise (or Hadamard) product, then we get time spent by residents at home.

We also define a matrix $\beta$ that describes how bites are distributed among humans:

$$\beta = \Theta^T \cdot \text{diag}\left(\frac{1}{W}\right)$$

Now, for example, we can define the number of bites, per person, per day, by strata. While $fqZ$ describes biting, and $fqZ/W$ is a measure of risk, with units bites per person, per day; $E$ computes bites, per person, per day summing risk over habitats weighted by time spent:

$$E = \Theta^T \cdot \text{diag}\left(\frac{fqZ}{W}\right) = \beta \cdot fqZ$$

**Vectorize** A lot of the hard work has been done in $\mathcal{B}$. As before, we rewrite

$$X = \left[\begin{array}{c} X_1 \\ X_2 \\ X_3 \\ X_4 \end{array}\right] \tag{23}$$

and we also transform the parameter

Now, we can rewrite the dynamics of infection as:

$$\frac{dX}{dt} = b\beta \cdot fqZ(H - X) - rX \tag{24}$$

and

$$\kappa = \beta^T \cdot cX \tag{25}$$

**Generic** If we take a birds-eye view of model building, we can describe model specification as an instance of a generic process:

- Define the model structure
  - Define the stratum membership matrix, $\mathcal{J}$.
  - Define the population strata, including the number of strata $n$ and the population size of each stratum, $H$.
  - Define a model of mobility, $\Theta$, that describes time spent.
  - Transform time spent into a mixing matrix, $\beta$, through a model of blood feeding, $\mathcal{B}$.
  - Define model inputs from $\mathcal{Y}$. Blood feeding among patches, at the net rate $fqZ$ is transformed into a measure of exposure distributed among patches as:

$$E = \beta \cdot fqZ$$

- Define the dynamics, $dX/dt$.
  - Define the state space. In this case, the model variables are fully defined by a column vector of length $n$: the density of infected humans in each stratum, $X$.
  - Define model parameters. In this case, $r$, $b$, and $c$.
  - The dynamics are described by a system of ordinary differential equations:

$$\frac{dX}{dt} = b\beta \cdot fqZ(H - X) - rX \tag{26}$$

- Define the outputs of $\mathcal{X}$
  - The primary output is defined by the probability a mosquito would become infected after biting a human in each stratum, $x$. This is defined by a relationship: $x = cXH$, so that $xH = cX$.
  - The NI is

$$\kappa = \beta^T \cdot cX$$

### Modular

After rewriting the model in modular notation, every parameter, variable, or term is either a vector or a matrix.

#### Habitat Structure

We assume the habitats are situated in in patches as defined by the habitat membership matrix $\mathcal{N}$, where the $\mathcal{N}_{i,j} \in \mathcal{N} = 1$ if the $i^{th}$ habitat is found in the $j^{th}$ patch. We let $\xi$ be a vector that describes the proportion of eggs laid in each habitat by the adult mosquitoes in that habitat, so

$$\mathcal{N}^T \cdot \xi = 1.$$

The egg laying matrix $\mathcal{U}$ is defined by

$$\mathcal{U} = \mathcal{N}^T \cdot \text{diag}(\xi)$$

Egg laying rates, $\Gamma$, are computed by $\mathcal{M}$, passed to $\mathcal{L}$, and transformed:

$$\eta = \mathcal{U} \cdot \Gamma \tag{27}$$

#### Aquatic Dynamics

We let $L$, a vector, denote larval densities in an aquatic population. Similarly, we let $\psi$ denote the maturation rate, $\phi$ the density independent mortality rate, and $\theta L$ the density dependent increase in mortality rates in response to mean crowding. The aquatic dynamics are described by the equations:

$$\frac{dL}{dt} = \eta - (\psi + \phi + \theta L)L \tag{28}$$

where every quantity in these equations is a vector of length $l$.

Emergence rates from the aquatic habitats, $\alpha$, are defined and transformed to emergence rates in patches, $\Lambda$, using the following pair of relationships:

$$\begin{array}{ll} \alpha & = \frac{\psi L}{2} \\ \Lambda & = \mathcal{N} \cdot \alpha \end{array} \tag{29}$$

The term $\Lambda$ is used in $dM/dt$.

#### Adult Mosquito Dynamics

We let $M$ denote adult mosquito density; $G$ gravid mosquitoes, and $Y$ infected mosquitoes. We let $g$ denote per-capita mortality in the patches, $\sigma$ the emigration rate, and $\mathcal{K}$ the dispersal matrix. From these, we define the demographic matrix:

$$\Omega = \text{diag}(g) + (I - \mathcal{K}) \cdot \text{diag}(\sigma)$$

We also let $f$ denote the overall blood feeding rate, $q$ the human fraction, $\gamma$ the egg laying rate. Given $\Lambda$ (from above) and $\kappa$ (defined below) equations describing adult mosquito ecology and infection dynamics are:

$$\begin{array}{ll} \frac{dM}{dt} & = \Lambda - \Omega \cdot M \\ \frac{dG}{dt} & = f(M - G) - \nu G - \Omega \cdot G \\ \frac{dY}{dt} & = fq\kappa(M - Y) - \Omega \cdot Y \end{array} \tag{30}$$

The core outputs of this dynamical component are egg laying rates:

$$\Gamma = \chi \nu G \tag{31}$$

and the density of infectious mosquitoes:

$$Z = e^{-\Omega\tau} Y \tag{32}$$

**Blood Feeding**

We formulate a matrix describing, $\Theta$, where $\Theta_{i,j} \in \Theta$ denotes the fraction of time spent by the $j^{th}$ population stratum in the $i^{th}$ patch. We let $H$ describe population density of each stratum. The density of humans in each patch is

$$W = \Theta \cdot H.$$

and we define $\beta$ as the mixing matrix:

$$\beta = \Theta^T \cdot \text{diag}\left(\frac{1}{W}\right)$$

**Parasite Infection Dynamics in Humans**

We let $X$ denote the density of infected and infectious humans. Let $r$ denote the rate that infections clear; $b$ the probability a mosquito becomes infected, per infectious bite; and $c$ the probability a mosquito becomes infected after blood feeding on a human in each stratum. Now, the dynamics of infection in the strata are described by:

$$\frac{dX}{dt} = b\beta \cdot fqZ \cdot (H - X) - rX \tag{33}$$

The primary output is the NI ($\kappa$). To compute NI, we first define the probability a mosquito becomes infected after biting an individual in each stratum is defined by $x = cX/H$. From this, we compute NI ($\kappa$)

$$\kappa = \beta^T \cdot cX = \beta^T \cdot xH \tag{34}$$
