## Supplement 4 - Vector Dynamics for "Spatial Dynamics of Malaria Transmission"

### Vector Dynamics

In the following, we:

- Present a dispersal matrix, $\mathcal{K}$, written out in long form.
- Solve for the steady states of the mosquito models presented in the main manuscript.
- Discuss extending a model to have multiple species, including a next-generation matrix for multi-vector models.
- Briefly discuss the problem of modeling immigrating mosquitoes.

#### The Dispersal Matrix

We assume mosquitoes move without retaining any memory of their position on a landscape: the resulting model is classified as Eulerian [1]. This framework does not assume, *a priori*, that the patches are gridded or that all mosquitoes move to a nearest neighbor, but gridded population models with nearest neighbor diffusion are a special case of this class of models. The underlying landscapes are allowed to be as heterogeneous with respect to adult movement as needed.

Let $\mathcal{K}(t)$ describe the fraction of mosquitoes emigrating from each patch that land on another patch: $\mathcal{K}_{i,j}(t) \in \mathcal{K}(t)$ is the fraction of mosquitoes leaving patch $j$ that end up in patch $i$. Each column in the matrix thus accounts for mosquitoes leaving from $j$:

$$\mathcal{K} = \left[\begin{array}{ccccc} j=1 & j=2 & j=3 & \dots & j=p \\ \boxed{\begin{array}{c} 0 \\ \mathcal{K}_{2,1} \\ \mathcal{K}_{3,1} \\ \vdots \\ \mathcal{K}_{p,1} \end{array}} & \boxed{\begin{array}{c} \mathcal{K}_{1,2} \\ 0 \\ \mathcal{K}_{3,2} \\ \vdots \\ \mathcal{K}_{p,2} \end{array}} & \boxed{\begin{array}{c} \mathcal{K}_{1,3} \\ \mathcal{K}_{3,2} \\ 0 \\ \vdots \\ \mathcal{K}_{p,3} \end{array}} & \boxed{\begin{array}{c} \dots \\ \dots \\ \dots \\ \ddots \\ \dots \end{array}} & \boxed{\begin{array}{c} \mathcal{K}_{1,p} \\ \mathcal{K}_{2,p} \\ \mathcal{K}_{3,p} \\ \vdots \\ 0 \end{array}} \end{array}\right] \quad (1)$$

The diagonal elements of $\mathcal{K}$ are all zeros – $\mathcal{K}$ is defined by the act of leaving. Just as emigration is related to resource availability, it may be true that the elements of $\mathcal{K}$ could be related to resource availability in the destination patch $i$. We also explicitly consider mortality and population loss from migration to areas outside the spatial domain associated with emigration from $j$. Let $\mu$ denote a vector describing the fraction of mosquitoes that die as a result of emigrating from a patch. A distance and resource-based approaches to mosquito movement is thus:

$$\mathcal{K}_{i,j} = \frac{(1-\mu_j) F_K(\mathcal{D}_{i,j}, B_j, Q_j, S_j)}{\sum_j F_K(\mathcal{D}_{i,j}, B_j, Q_j, S_j)}.$$

After computing $\sigma$ (Eq. 19) and $\mathcal{K}$, mosquito demography, $\Omega$ can be computed with Eq. 20.

### Steady States

We can solve for steady state relationships within each one of the components. These can be put together to arrive at the steady state for the entire system.

**Adult Mosquitoes** We consider mosquito population dynamics at the steady state as a forced system with two inputs, $\Lambda$, and $\kappa$.

At the steady state, adult mosquito density is:

$$M = \Omega^{-1} \cdot \Lambda. \tag{2}$$

Because the density of gravid mosquitoes does not depend on infection status in this model, it can be solved at equilibrium given $M$. The diagonal matrices arise because $\nu G = \text{diag}(\nu) \cdot G$, which is needed in order to factor $G$ out of the equation and solve.

$$G = (\text{diag}(\nu) + \Omega + \text{diag}(f))^{-1} \cdot \text{diag}(f) \cdot M \tag{3}$$

We can formulate a model for the fraction parous (bloodfed), $V$, where

$$\frac{dV}{dt} = fq(M - V) - \Omega \cdot V \tag{4}$$

At the steady state, the fraction parous is:

$$V = (\text{diag}(fq) + \Omega)^{-1} \cdot \text{diag}(fq) \cdot \Omega^{-1} \cdot \Lambda. \tag{5}$$

Similarly, the density of infected mosquitoes is:

$$Y = (\text{diag}(fq\kappa) + \Omega)^{-1} \cdot \text{diag}(fq\kappa) \cdot \Omega^{-1} \cdot \Lambda. \tag{6}$$

Under static conditions, $\Upsilon_\tau = e^{-\Omega\tau}$, and at the steady state,

$$fq\kappa(M - Y) = f_\tau q_\tau \kappa_\tau (M_\tau - Y_\tau) = \Omega \cdot Y,$$

so we can make a substitution into Eq. 25, and the density of infective mosquitoes at the steady state is given by:

$$e^{-\Omega\tau} \cdot \Omega \cdot Y = \Omega \cdot Z. \tag{7}$$

so

$$Z = \Omega^{-1} \cdot e^{-\Omega\tau} \cdot \Omega \cdot Y. \tag{8}$$

or

$$Z = \Omega^{-1} \cdot e^{-\Omega\tau} \cdot \Omega \cdot (\text{diag}(fq\kappa) + \Omega)^{-1} \cdot \text{diag}(fq\kappa) \cdot \Omega^{-1} \cdot \Lambda. \tag{9}$$

Note that in the limit as $\kappa$ approaches zero, that

$$\begin{aligned} \mathcal{V} &= \lim_{\kappa \to 0} \tfrac{fqZ}{W} \\ &= \lim_{\kappa \to 0} fq\Omega^{-1} \cdot e^{-\Omega\tau} \cdot \Omega \cdot (\text{diag}\,(fq\kappa) + \Omega)^{-1} \cdot \tfrac{fq\kappa M}{W} \\ &= fq\Omega^{-1} \cdot e^{-\Omega\tau} \cdot \text{diag}\left(\tfrac{fqM}{W}\right). \end{aligned} \tag{10}$$

**Estimation** In practical analysis of surveillance data from malaria or other mosquito-borne pathogens, it is common to get incidence or prevalence data from the human population, from which an estimate of $Z$, the density of infectious (sporozoite positive) mosquitoes can be obtained.

Noting that $(e^{-\Omega\tau})^{-1} = e^{\Omega\tau}$, we can solve for Y:

$$Y = \Omega^{-1} \cdot e^{\Omega\tau} \cdot \Omega \cdot Z. \tag{11}$$

and

$$M = \text{diag}\left(\frac{1}{fq\kappa}\right) \cdot \text{diag}\,(fq\kappa + \Omega) \cdot Y \tag{12}$$

and

$$\Lambda = \Omega \cdot M \tag{13}$$

**Aquatic Mosquitoes** The simplest nonlinear model of aquatic development including density-dependent mortality was given as:

$$\dot{L} = \eta - (\psi + \phi + \theta L)L \tag{14}$$

Given $G$, the rate that eggs are deposited into habitats, $\eta$ is known. Additionally, we assume $\Lambda$ is known so that we can solve for $L$:

$$L = \Lambda/\psi \tag{15}$$

From this we can solve for the density-dependent term:

$$\theta = (\eta - \psi L - \phi L)/L^2 \tag{16}$$

### Multiple Vector Species

To formulate models with multiple mosquitoes-species, we let $s$ denote the number of vector species. We use different relative daily activity rates ($\xi$), biting weights ($w_f$), and weighted availability of alternative hosts ($B$ and $\zeta$) to modify the same TiSp matrix, $\Theta$. Each species would also have its own functional response, so that the TaR matrix ($\Psi$), host availability ($W$), biting distribution matrix ($\beta$), human biting habit ($q$), and feeding rates ($f$) would also be species-specific.

These multi-species models also draw attention to differences among vector species in their ability to host the parasite. The notion of vectorial capacity makes the assumption that a host is *perfectly infectious* so that the formulas focus on phenomena related to the vectors, but here we define *vector competence* as differences in the fraction of mosquitoes that would become infected and remain infected through sporogony, assuming the mosquitoes survived. We let $c_j$ denote this fraction, for the $j^{th}$ species.

Parasite next-generation matrices for models in which there are two or more vector species are slightly more complex. Since the TaR matrix could be different for the same humans, depending on mosquito activity patterns, we let $\Psi_i$ be the TaR matrix for the $i^{th}$ vector and we let $\mathcal{D}_i$ be the associated HTC matrix (accounting for differences in mosquito preferences and time at risk), and we let $\mathcal{V}_i$ denote the vectorial capacity matrix for the $i^{th}$ species.

$$R_{Z_i} = b\beta_i \cdot f_i q_i \Omega_i^{-1}. \tag{17}$$

How many infective mosquitoes would arise from each human infection? The answer is $s$ matrices, of dimension $p \times n$, describing transmission from a human in each stratum to mosquitoes of each species in each patch:

$$R_{X_j} = c_j e^{-\Omega_j \tau} \cdot f_j q_j M_j \cdot \beta_j^T \cdot \text{diag}\left(DH\right). \tag{18}$$

The types next generation with multiple vector species is an $n + ps \times n + ps$ block matrix:

$$\mathcal{N} = \left[\begin{array}{ccccc} 0 & R_{X_1} & R_{X_2} & \dots & R_{X_s} \\ R_{Z_1} & 0 & 0 & \dots & 0 \\ R_{Z_2} & 0 & 0 & \dots & 0 \\ \vdots & \vdots & \vdots & \ddots & \vdots \\ R_{Z_s} & 0 & 0 & \dots & 0 \end{array}\right] \tag{19}$$

When we square this, we get a matrix in block form:

$$\mathcal{N}^2 = \left[\begin{array}{cc} \mathcal{R} & 0 \\ 0 & \mathcal{Z} \end{array}\right] \tag{20}$$

Transmission from humans to humans through each vector species, $\mathcal{R}_i$, where $\mathcal{R}$ is a $n \times n$ block matrix

$$\mathcal{R} = \sum_i b\beta_i \cdot \mathcal{V}_i \cdot \beta_i^T \cdot \text{diag}(DH) \tag{21}$$

and $\mathcal{Z}$ is a $ps \times ps$ matrix with the block form:

$$\mathcal{Z} = \left[\begin{array}{cccc} \mathcal{Z}_{1,1} & \mathcal{Z}_{1,2} & \dots & \mathcal{Z}_{1,s} \\ \mathcal{Z}_{2,1} & \mathcal{Z}_{2,2} & \dots & \mathcal{Z}_{2,s} \\ \vdots & \vdots & \ddots & \vdots \\ \mathcal{Z}_{s,1} & \mathcal{Z}_{s,2} & \dots & \mathcal{Z}_{s,s} \end{array}\right] \tag{22}$$

where each block matrix

$$\mathcal{Z}_{i,j} = R_{X_j} \cdot R_{Z_i}$$

is a $p \times p$ matrix that describes parasite transmission from the $i^{th}$ mosquito species populations in the patches through human strata back to the $j^{th}$ mosquito species in the patches.

#### Immigrating Mosquitoes

We can add malaria importation in mosquitoes by simply adding a term:

$$\frac{dM}{dt} = \Lambda - \Omega \cdot M + \delta_M \tag{23}$$

Importation of infected mosquitoes is highly problematic because of the time delay for the EIP, so we ignore immigrating infected mosquitoes, and count them only if they survive to become infective. Importation of infective mosquitoes is:

$$\frac{dZ}{dt} = \Upsilon_\tau \cdot \text{diag}(f_\tau q_\tau \kappa_\tau) \cdot (M_\tau - Y_\tau) - \Omega \cdot Z + \delta_Z \tag{24}$$
