## Supplement 5 - Human Mobility for "Spatial Dynamics of Malaria Transmission"

*Supplement 5 - Human Travel and Mobility*

#### Human Mobility

In developing a framework for modeling parasite dispersal by moving humans, we were motivated by a concept of location and risk based on exposure patterns in sub-Saharan Africa, where exposure tends to occur indoors at night. The conceptual model for human travel we use is herein called the *domestic* modality for travel and mobility. In the domestic modality, people have a home, the place where they return home to sleep most nights. Time spent is subdivided into three classes: travel, which is defined by a night spent away from home; time spent at home; and mobility, which includes locations around home, not counting home or travel. In malaria, substantial research has focused on travel and malaria [1–3], particularly in malaria elimination settings [4]. Human mobility has been of greater interest in the study of dengue transmission dynamics [5, 6], and substantially less attention in malaria has been given to mobility patterns (but see [7]). Clearly, even areas where transmission mostly occurs at night, some biting occurs away from home during the day, or at dawn or dusk, when night biting mosquitoes are active [8, 9].

In developing a mathematical framework to model human travel and mobility based on the domestic modality, we assume there is some underlying model describing humans' locations as they move around inside a spatial domain [10, 11]. We assume mobility within the spatial domain is occurring over comparatively fast time scales, as people move around the area every day for work, school, shopping, church, or various other activities, perhaps even a short overnight trip to an urban center, or to visit relatives. We further assume exposure to the bites of infective mosquitoes at a place is roughly proportional to the amount of time spent there, and since the generation time of malaria is quite long – the shortest possible serial interval for malaria is approximately 25-30 days – we have formulated a model in terms of the average time spent among a set of patches [10, 12]. For diseases with shorter generation times, it may be useful to model the dynamics of human travel explicitly.

The domestic modality could apply to other sub-populations whose travel patterns differ from the defined type. Obviously, there are some people who have multiple homes, or who spend their nights at work and sleep during the day. In this framework, travel and mobility by these sub-populations would differ from the rest of the population, so they could be segmented into a separate stratum. With some care, the same framework might be adaptable to model transmission in populations with other modalities for travel and mobility:

- *Nomadism* is defined for populations that move around without establishing a home.

- *Migration* includes all mobility related to moving a household, permanently changing a home base.
- *Seasonal Labor Migration* is defined for populations that have two or more homes that they use in an alternating pattern at different times of the year.
- *Forest Malaria* is a modality that applies to populations where there is a comparatively low risk of malaria at home, but the risk of malaria is high for travel or mobility away from home. In models of forest malaria, steady state assumptions are likely to give a misleading picture.

The list is far from exhaustive, and there may be many situations where travel and mobility shape transmission in some particular ways that are important for both the epidemiology and control.

#### Simple Trip

One way to develop a time spent for mobility in the domestic modality is by modifying the "simple trip model," which describes how hosts travel temporarily to other locations before returning home [10, 13]. Here, we define the model for time spent by a single stratum.

Let $H_j$ denote the density of humans in the $j^{th}$ patch who reside in the $i^{th}$ patch, out of a total of $p$ patches, where they are at risk. We also consider how much time humans spend in places where they are not at risk, so we add two patches: time spent traveling, and time spent in places where there is no risk. Time is spent either at home or away in a sequence of trips:

$$\begin{aligned} \frac{dH_i}{dt} &= -\sum_{j=1}^{p+2} \phi_j H_i + \sum_{j=1}^{p+2} \tau_j H_j \\ \frac{dH_j}{dt} &= -\tau_j H_j + \phi_i H_i \end{aligned} \tag{1}$$

The constant $\phi_j$ represents the rate at which hosts travel to $j$, the constant $\tau_j$ is the rate at which hosts visiting $j$ return home, and we assume $\phi_i = \tau_i = 0$.

When the movement equations reach a steady state and the derivatives on the left hand side of Eq. 1 equal zero, the population $H_i$ is distributed across the $p$ metapopulation sites, travel, and not at risk, as follows:

$$\begin{aligned} \theta_i^* &= \frac{1}{1 + \sum_{j=1}^{p+2} \frac{\phi_j}{\tau_j}} \\ \theta_j^* &= \frac{\phi_j}{\tau_j} \frac{1}{1 + \sum_{j=1}^{p+2} \frac{\phi_j}{\tau_j}} \end{aligned}$$

These $\theta$ describe the fraction of time spent in each patch, and $\sum_{j=1}^{p+2} = 1$

Let $d$ denote the time of day, $d = t - \text{floor}(d)$. We note that this model would not output time spent by time of day unless the parameters depended on time of day, $\phi_j(d)$ and $\tau_j(d)$. In the resulting model, we would need to numerically solve Eqs. 1 to get $\theta(d)$.

Let $\Theta$ denote the time spent matrix. A time spent matrix is constructed by considering the travel patterns of each population stratum. Each column in $\Theta$ represents the first $p$ elements of a time spent vector, $\theta_j$ where $j \in 1, 2, \ldots, p$. By design, the time spent matrix does not include time spent during travel or not at risk:

$$\Theta = \left[ \begin{array}{ccccc} i=1 & i=2 & i=3 & \ldots & i=n \\ \boxed{\begin{array}{c} \Theta_{1,1} \\ \Theta_{2,1} \\ \Theta_{3,1} \\ \vdots \\ \Theta_{p,1} \end{array}} & \boxed{\begin{array}{c} \Theta_{1,2} \\ \Theta_{2,2} \\ \Theta_{3,2} \\ \vdots \\ \Theta_{p,2} \end{array}} & \boxed{\begin{array}{c} \Theta_{1,3} \\ \Theta_{2,3} \\ \Theta_{3,3} \\ \vdots \\ \Theta_{p,3} \end{array}} & \boxed{\begin{array}{c} \ldots \\ \ldots \\ \ldots \\ \ddots \\ \ldots \end{array}} & \boxed{\begin{array}{c} \Theta_{1,n} \\ \Theta_{2,n} \\ \Theta_{3,n} \\ \vdots \\ \Theta_{p,n} \end{array}} \end{array} \right] \tag{2}$$

The rows should sum up to less than one, if humans travel at all, or if they spend any time in locations where they are not at risk, such as automobiles, or office buildings.

These time-spent matrices are designed for short-term projections, so population changes due to human births, deaths, and migration are either assumed to be small, or they are dealt with through stratification. The underlying assumption is that over short time scales, daily mobility patterns play a dominant role in moving parasites around within the spatial domain. Malaria importation by residents occurs during travel, or time spent outside of the spatial domain (Supplement 4). The concept of residency is flexible, but a residence is where a person would be counted in a census. Those whose are only present for a short time are "visitors", and we use the concept of a visitor population to model a secondary malaria immigration route (Supplement 4).

**Time Spent *vs.* Distance** While the "simple trip" model helps to clarify how time spent *could* be computed, an alternative way of generating parameters for time-spent matrices is the following:

- Let $p_i$ denote the fraction of time spent in the home patch, $i$.
- Let $p_t$ denote the fraction of time spent traveling or not at risk.
- Let $\theta$ denote time spent, where $\theta_i = p_i$ and $\theta_{j \neq i}$ be a function of distance and perhaps some features of the destination patch $j$, and $\sum_{j \neq i} \theta_j = 1 - p_i - p_t$.

The functions describing time spent by distance and features of a destination can be informed by data.

**Time at Risk** Models of blood feeding must thus merge time spent, $\Theta(t)$, as defined above, and mosquito activity rates. From this synthesis, we define three related concepts: a matrix describing time at risk (TaR), a vector describing average daily mosquito blood feeding rates $f$ in each patch, and a vector describing the proportion of blood meals that are taken on humans in each patch, $Q$.

Let $\xi(d)$ describe relative daily blood feeding activity for a malaria vector species over a day.

In some cases, we can consider how $\xi$ varies over time, and we want functions such that:

$$\int_0^1 \xi(t)dt \approx 1 \tag{3}$$

If we measured mosquito activity patterns, then for some large integer value of $T \gg 1$,

$$\int_0^T \xi(t)dt \approx T \tag{4}$$

and

$$\xi(d) = \frac{1}{T}\sum_T \xi(d+T) \tag{5}$$

Note that this is defined in a similar way to the concept of average time spent over a day, $\Theta(t)$. We note that $\xi(t)$ describes blood feeding activity rates and *not* the distribution of blood meals, which should be thought of as the product of activity and host availability.

A *time at risk* (TaR) matrix, $\Psi$, is constructed from a time spent matrix. It is like $\Theta$ in every way, except that it weights time spent throughout the day by mosquito blood feeding activity rates. Notably, this implies that models with more than one mosquito species would weight time spent differently, so that while there is one time spent matrix, $\Theta$, there could be as many TaR matrices as vector species, with activity patterns defined by $\xi_i(d)$. The resulting species-specific TaR matrices would be:

$$\Psi_i = \int_0^1 \text{diag}\left(\xi_i(d)\right) \cdot \Theta(d)dt \tag{6}$$

### Malaria Importation

**The Travel FoI** Here, we define malaria risk during activities related to travel, where at least one night is spent away from home. Travel malaria is often defined by a difference in prevalence of malaria associated with recent travel.

Data on travel is often available form Malaria Indicator Surveys and from Demographic Health Surveys, reported as recent travel, or travel within the last $s$ days. Let $T$ denote the fraction of a population that is traveling, $\alpha$ the frequency of travel, and $1/\lambda$ the length of a trip:

$$\frac{dT}{dt} = \alpha(1-T) - \lambda T \tag{7}$$

So that the fraction of the population that is traveling is:

$$\hat{T} = \frac{\alpha}{\alpha+\lambda} \tag{8}$$

Let $\sigma = s^{-1}$. The fraction that has recently traveled is modeled as:

$$\frac{dS}{dt} = \lambda T - \sigma S \tag{9}$$

It follows that

$$S = \lambda s T = \frac{s\alpha(\lambda-\alpha)}{\alpha+\lambda} \tag{10}$$

If we let $\hat{h}$ denote the average FoI while traveling. We define the travel FoI as:

$$\delta = \hat{h}\hat{T} \tag{11}$$

**Visitors** The concept of a "visitor" population is a model choice. A "visitor" population is present with parameters unaffected by local transmission dynamics. With immigrant labor, it might make more sense to create a resident population with frequent travel to the mainland.

The visitor population is modeled as an available population, a weighted, ambient population at risk in each patch, $W_\delta$. In some cases, it may be useful to explicitly model an ambient visitor population $A_\delta$ before assigning it a weight, so that it can be compared directly with $A_r$ or $A_n$.
